# A Two-Stage Questionnaire and Actigraphy Screening for Isolated REM Sleep Behavior Disorder in a Multicenter Cohort

**DOI:** 10.64898/2026.03.09.26347106

**Authors:** Caleb A. Massimi, Giorgio Ricciardiello Mejia, Andre Metzger, Kang Hyun Ryu, Salonee Marwaha, Eva Grzegorczyk, Li Zhou, Eliana Jacobs, Boris Gilyadov, Claudia Kunney, Liqhwa Ncube, Ankit Parekh, Emmanuel Mignot, Fanny M Elahi, Joseph Winer, Kathleen Poston, Andreas Brink-Kjaer, Emmanuel H. During

**Affiliations:** Department of Neurology, Icahn School of Medicine, New York, USA; Department of Medicine, Division of Pulmonary, Critical Care and Sleep Medicine, Icahn School of Medicine, New York, USA; Department of Artificial Intelligence and Human Health, Icahn School of Medicine, New York, USA; Department of Psychiatry, Division of Sleep Medicine, Stanford University, USA; Department of Neurology, Stanford University, Palo Alto, USA; Department of Health Technology, Technical University of Denmark, Kongens Lyngby, Denmark

## Abstract

**Objective:** Isolated rapid-eye-movement sleep behavior disorder is a prodromal marker of synucleinopathies. However, most cases remain undiagnosed due to the insufficient predictive value of questionnaires and limited access to confirmatory video-polysomnography. We assessed a two-stage screening strategy combining a brief questionnaire on rapid-eye-movement sleep behavior disorder symptoms and other prodromes with wrist actigraphy across multiple case-control cohorts.

**Methods:** Participants aged 40–80 without neurodegenerative disease were recruited from five cohorts; all cases were confirmed by video-polysomnography. The questionnaire was administered to 289 participants, and 236 underwent ≥14 nights of home wrist actigraphy. The wearable-based algorithm was built on four movement features (mean motor activity, activity index, short or long immobile bouts, twitch activity). Models were trained with nested cross-validation using XGBoost.

**Results:** The full retrospective cohort included 396 participants (99 cases, 297 controls; mean age 64 ± 11; 55% male). The dream enactment question alone achieved an area under the curve of 0.85, which improved to 0.86 using the four-item questionnaire. Actigraphy alone achieved 82% sensitivity and 84% specificity. In the subgroup completing both assessments (75 cases, 54 controls), the two-stage protocol—questionnaire followed by actigraphy—yielded 68% sensitivity and 100% specificity using the dream-enactment question alone, and 73% sensitivity and 100% specificity using the four-item questionnaire.

**Interpretation:** A two-stage protocol combining questionnaire and actigraphy demonstrated high specificity and good sensitivity for detecting isolated rapid-eye-movement sleep behavior disorder in this multicenter cohort. This low-cost, scalable strategy is compatible with widely used wearable devices and warrants validation in community-based populations.

## Introduction

Isolated (or idiopathic) REM sleep behavior disorder (iRBD) is a remarkably specific prodromal marker of synucleinopathies, such as Parkinson’s disease (PD) or dementia with Lewy bodies (DLB).^1,2^ Longitudinal studies indicate that most individuals with iRBD (>80%) will eventually develop a synucleinopathy, making it a robust predictor of future neurodegeneration.^3,4^ RBD manifests as abnormal movements during REM sleep, ranging from brief twitches to overt dream enactment, and is commonly associated with other prodromal features such as hyposmia, constipation, and orthostatic symptoms. Although iRBD affects more than 1% of people over age 50,^1,5–8^ most patients remain undiagnosed due to two main limitations: (1) In-laboratory video-polysomnogram (vPSG), the gold standard for definitive diagnosis, is costly, difficult to interpret^9^, and likely will never be widely available^10^; (2) Questionnaires focusing on RBD symptoms (e.g., dream enactment) have shown a low positive predictive value (∼10%) in community-based samples, limiting their utility as standalone screening tools.^11–13^ Staged protocols (sequential questionnaires followed by clinical evaluation) remain resource-intensive and imprecise, leading to erroneous diagnosis in at least one-third of cases.^14–17^ This “diagnostic bottleneck” has kept RBD cohorts small and possibly biased towards severe cases, which may not represent the broader iRBD population.

Wrist-worn wearables have emerged as promising tools for scalable, low-cost screening of RBD. We and others have developed machine-learning (ML) models based on wrist accelerometer data^18,19^ achieving high diagnostic performance (AUC ranging 0.84-0.97) across 6 independent cohorts and devices,^18,20,21^ demonstrating robust generalization. We previously demonstrated that such models, relying on interpretable actigraphy features, could be fully automated without requiring manual sleep diary input or sleep-wake annotations.^21^ Although algorithmic performance will likely continue to improve, wrist-based actigraphy and questionnaires are unlikely to function as standalone screening tools given the low prevalence of iRBD.

In prior work, we observed that pre-selecting participants using a single-item question about dream enactment improved the specificity of a two-stage screening approach combining questionnaire and actigraphy.^18^ Similarly, enrichment based on additional prodromal features such as hyposmia, constipation, or orthostatic symptoms further increased precision.^18,20^ These findings motivated the hypothesis that an augmented, multi-prodrome RBD screening questionnaire (Stage 1), followed by actigraphy (Stage 2), could provide a scalable framework with improved precision compared to single-item pre-screening, while maintaining adequate sensitivity.

This study pursued three objectives: (1) to evaluate a four-item prodromal questionnaire for iRBD in a large retrospective, multicenter case–control study, recruited from sleep clinics, and to compare the performance of individual items with that of a ML model trained on all prodromal features; (2) to train and validate a wearable-based detection model in a larger and more diverse dataset than in prior studies; and (3) to assess a sequential two-stage screening strategy—questionnaire followed by actigraphy—in preparation for community-based studies.

## Methods

### Study Participants and Ethical Approval

This study was approved by the Institutional Review Boards of the Icahn School of Medicine at Mount Sinai and Stanford University, and all research participants provided written informed consent. Inclusion criteria were an age between 40 and 80 years and the absence of an overt neurodegenerative disease. All RBD diagnoses were confirmed by overnight video-polysomnography (vPSG) according to ICSD-3 criteria.^22^ Control participants required the absence of dream-enactment symptoms or demonstrated normal REM sleep atonia on vPSG. All available participants meeting these criteria were included; no individuals were excluded based on questionnaire responses or actigraphy outcomes. Neurodegenerative diseases were excluded through clinical history, electronic medical record review, and clinician assessment at each study site.

Mount Sinai research cohorts included the Sleep and Healthy Aging Study (SHAS; n = 62) and the VascBrain cohort (n = 25), which provided questionnaire and wearable data (**Table 1**). SHAS and VascBrain are longitudinal cohorts enrolling adults with or without RBD to study neurodegenerative disease. At Mount Sinai, additional data were collected in 147 consecutive patients in sleep medicine clinics, where the questionnaire was integrated into standard-of-care assessments.

**Table 1.** Demographic distributions of participants stratified by diagnosis and by cohort. Percentages are based on non-missing values. Statistically significant between-cohort differences were observed for BMI (p-value=0.0039), Gender (p-value=0.0007) and Race (0.0125). When stratified by cohort, VascBrain patients were younger 55.7±9.1 years on average.

|  | Diagnosis |  |  |  |  |
| --- | --- | --- | --- | --- | --- |
|  | iRBD<br>N=99 | Control<br>N=297 | p-value |  |  |
| Age [years] | 66.6 ± 8.7<br>(99) | 63.7 ± 10.9<br>(296) | 0.0525 |  |  |
| BMI [kg/m²] | 26.3 ± 4.3<br>(99) | 28.6 ± 6.4<br>(206) | 0.0039 |  |  |
| Gender (Male) | 69 (69.7%) | 149 (50.2%) | 0.0007 |  |  |
| Race | -- | -- | 0.0125 |  |  |
| White | 74 (80.4%) | 194 (68.5%) | - |  |  |
| Black or African American | 8 (8.7%) | 23 (8.1%) | - |  |  |
| Asian | 5 (5.4%) | 16 (5.6%) | - |  |  |
| Other | 5 (5.4%) | 45 (15.9%) | - |  |  |
| Mixed | 0 (0.0%) | 5 (1.8%) | - |  |  |
|  | Cohort |  |  |  |  |
|  | Sinai<br>N=147 | SHAS<br>N=62 | Stanford<br>N=84 | VascBrain<br>N=25 | ADRC<br>N=78 |
| Age [years] | 60.7 ± 11.1<br>(147) | 65.8 ± 9.4<br>(61) | 65.8 ± 8.8<br>(84) | 55.7 ± 9.1<br>(25) | 71.7 ± 7.0(78) |
| BMI [kg/m²] | 29.8 ± 6.6<br>(147) | 25.7 ± 4.6<br>(60) | 26.1 ± 4.4<br>(84) | 26.9 ± 4.0<br>(14) | - |
| Gender (Male) | 75 (51.02%) | 37 (59.7%) | 62<br>(73.81%) | 10<br>(40.0%) | 34<br>(43.59%) |
| Race | -- | -- | -- | -- | -- |
| White | 69 (50.0%) | 35 (66.1%) | 77<br>(91.67%) | 19<br>(86.36%) | 68<br>(87.18%) |
| Black or African American | 21 (15.22%) | 8 (15.09%) | 1 (1.19%) | 0 (0.0%) | 1 (1.28%) |
| Asian | 6 (4.3%) | 2 (3.8%) | 6 (7.1%) | 0 (0.0%) | 7 (9.0%) |
| Other | 41 (29.7%) | 7 (13.2%) | 0 (0.0%) | 1 (4.5%) | 1 (1.3%) |
| Mixed | 1 (0.7%) | 1 (1.9%) | 0 (0.0%) | 2 (9.1%) | 1 (1.3%) |
| iRBD | 12 (8.2%) | 44 (71%) | 42 (50%) | 1 (4%) | 0 (0%) |
| Questionnaire (yes) | 147 (100) | 58 (93.5%) | 84 (100%) | 25 (100) | 0 (0%) |
| Actigraphy (yes) | 0 (0.0%) | 49 (79.03%) | 84<br>(100.0%) | 25<br>(100.0%) | 78<br>(100.0%) |

At Stanford, the Sleep Center cohort (n = 84) contributed questionnaire and wearable data, primarily from clinic participants (n = 63) and from community volunteers (n = 21) enrolled in a research study on RBD.^18^ The Stanford Alzheimer’s Disease Research Center (ADRC) contributed wearable data (n = 78). This retrospective multicenter case-control study adheres to the STROBE (Strengthening the Reporting of Observational Studies in Epidemiology) guidelines.

### Data Collection

#### Questionnaire

The four-item questionnaire screened for: (1) dream enactment using the validated RBD-Innsbruck summary question^22^, defined as kicking or hitting during sleep due to defensive dreaming; (2) Hyposmia, defined as reduced smell or taste compared to others or previous ability^18^; (3) Constipation, defined as requiring straining or laxative use^23^; (4) Orthostatic symptoms, suggestive of hypotension.^23^ Each item allowed three responses: *No* (0), *Don’t know* (0.5), or *Yes* (1). The dream enactment question was chosen based on simplicity and for its high specificity^22^ compared to other screening questionnaires.^14^ Questionnaire data were collected in 95 iRBD and 194 controls. No participants were excluded based on questionnaire response.

#### Wearable data

Participants wore a wrist-mounted triaxial accelerometer (AX3 or AX6, Axivity Ltd., Newcastle, UK) continuously on the dominant wrist. Sampling frequencies ranged from 25 to 100 Hz depending on cohort protocol.

#### Sleep detection

A total of 6,620 nights of raw recordings (including non-wear) were available from 78 cases with iRBD and 158 controls. In our research protocols, devices are typically shipped to participants and returned by mail, which can lead to extended periods (days to weeks) of non-wear. Accordingly, a key preprocessing step is to distinguish wear from non-wear nights. To this end, we implemented a previously validated automated night-wear detection pipeline ^21^ based on three criteria: (1) <4 hours of inferred night non-wear; (2) Total inferred sleep duration >3 hours and <12 hours; (3) Mean nightly skin temperature <27°C (suggesting device removal). These thresholds were defined a priori and applied identically across all participants and cohorts. After quality filtering, 3,819 nights were available for analysis, including 1,652 (mean 20.9 ± 8.8) nights in participants with iRBD, and 2,167 (mean 13.6 ± 5.4) in controls.

#### Actigraphy features

Building on our prior validated work^17^, we computed 113 candidate features derived from 1-second activity counts. This decision was motivated by evidence from our recent work indicating that rest–activity rhythm metrics show limited generalizability across cohorts (19), in contrast to sleep-related movement features.

Features were variations and time-window adaptations of four primary movement constructs capturing an excess of movement during sleep, described by others^24–27^–– ***Mean motor activity (MA)***: average activity in sleep; ***Activity index (AI)***: % epochs with any activity (varying epoch length x ∈ {10, 30, 60} s); ***Short and Long immobile bouts (SIB, LIB)***: zero-activity bouts of a certain duration/hour. SIB were defined as x ∈ {0, 1, 5} to 60 s. LIB were defined as > x ∈ {60, 120, 300} seconds. In addition, we included a novel feature developed by our group,^18^ ***Twitch activity (TA)***: number of non-zero activity counts with amplitude < x ∈ {0.5, 1, 1.5} g (gravity units) surrounded by zero-activity (**Figure 1**). Various time windows, including all night and the period starting 70 minutes and ending 200 minutes after predicted sleep onset––maximizing the proportion of REM sleep, and Hjorth parameters––capturing movement complexity, were applied, in addition to traditional features of sleep (e.g., wake after sleep onset, sleep efficiency).

**Figure 1:**
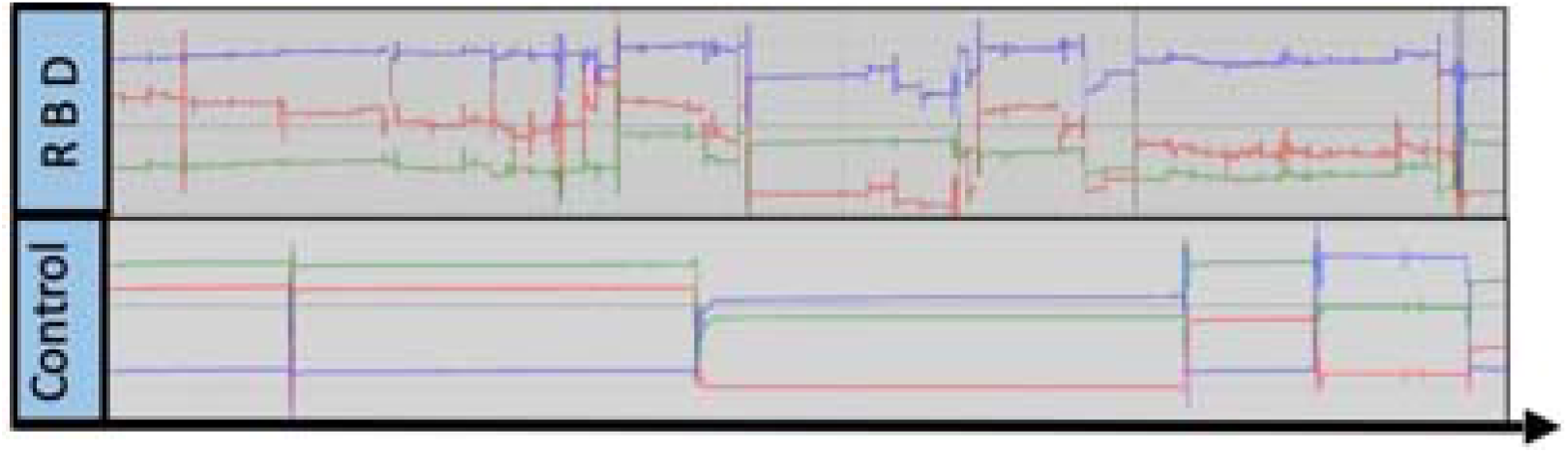
Wrist Accelerometer Signals During REM Sleep in an Individual with Isolated RBD and a Control. Representative raw tri-axial wrist accelerometer signals (x, y, z axes) recorded with the AX-6 device during a 40-minute segment of nocturnal REM sleep in an individual with isolated RBD and a control participant without RBD during an overnight video-polysomnogram. Although some movements are present in the control participant—likely reflecting occasional twitches, brief arousals, or comfort-related movements—the individual with RBD exhibits a higher frequency of sleep-related movements, including frequent low-amplitude activity consistent with twitching. Note that no overt dream-enactment behaviors were observed during this REM sleep cycle based on review of the synchronized video data.

**Figure 2:**
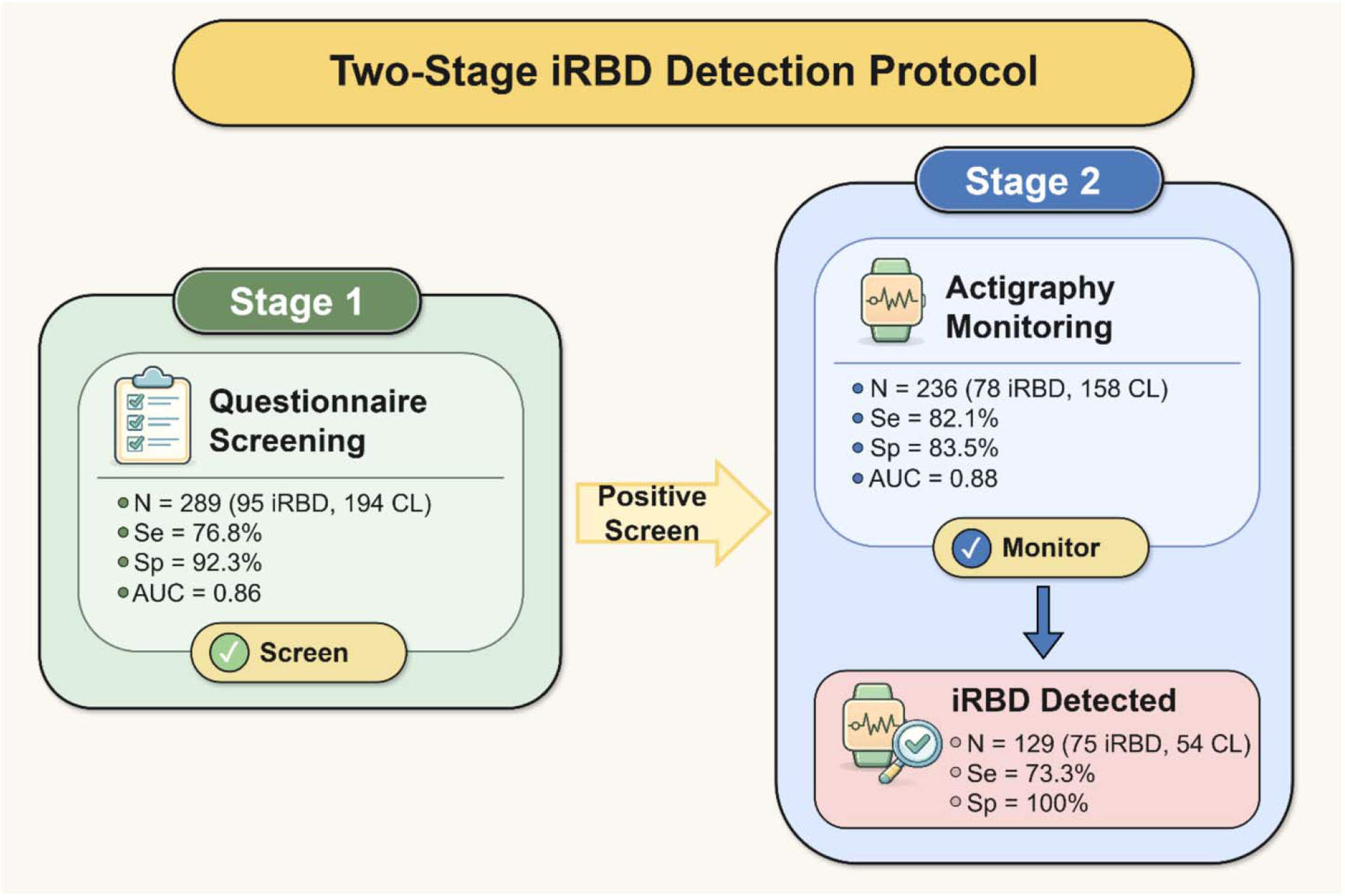
Two-Stage Protocol. Separate machine-learning classifiers were trained using questionnaire-based and actigraphy-derived features. For two-stage screening, participants were classified as having isolated REM sleep behavior disorder (iRBD) only if both classifiers yielded positive predictions, using a sensitivity-optimizing threshold for questionnaire screening and a specificity-optimizing threshold for actigraphy confirmation.

### Statistical Analysis

Descriptive statistics were calculated for questionnaire responses, demographics, and cohort characteristics. Continuous variables are presented as mean ± standard deviation (SD), and categorical variables as counts and percentages. Group comparisons were performed using Welch’s t-test or Mann–Whitney U test for continuous variables and Chi-square or Fisher’s exact test for categorical variables, as appropriate. Group comparisons were performed across three stratifications: (1) cohorts, (2) diagnosis (iRBD vs. controls), and (3) actigraphy availability. For cohort-level analyses, only descriptive statistics were shown.

### Questionnaire Modeling

To assess the association between questionnaire responses and iRBD diagnosis, logistic regression models were fitted using dummy-coded predictors with “No” as the reference category (Yes vs No; Don’t Know vs No). Three prespecified models were evaluated: (1) A full dummy-coded logistic regression including all Yes and Don’t Know terms; (2) A Don’t Know–only logistic regression to assess the independent contribution of uncertainty responses; (3) A combined model including all dummy-coded predictors.

Odds ratios (ORs), 95% confidence intervals (CIs), p-values, log-likelihood, Akaike information criterion (AIC), Bayesian information criterion (BIC), and McFadden’s pseudo-R^2^ were reported.

To explore potential higher-order interactions between questionnaire items, a penalized logistic regression model with L1 regularization (LASSO) was fitted. Polynomial interaction terms (interaction-only, up to degree 7) were generated and standardized prior to model fitting. Regularization strength was selected using internal 5-fold cross-validation within the dataset. Non-zero coefficients were extracted to identify informative main effects and interactions.

Performance metrics for the LASSO model—including ROC AUC, precision–recall AUC, Brier score, log loss, and pseudo-R^2^—were calculated on the full dataset and are reported descriptively to characterize model discrimination and calibration. These estimates were not used for final predictive evaluation, which relied on nested cross-validation as described below.

### Predictive Modeling

Predictive performance of questionnaire and actigraphy classifiers was evaluated using nested cross-validation. Performance metrics—including sensitivity, specificity, ROC AUC, and precision–recall AUC—were aggregated across outer folds. Ninety-five percent confidence intervals were estimated using the Student’s t-distribution across folds (α = 0.05).

Adjusted positive predictive value (PPV) was calculated assuming a population prevalence of 1.5%.

All statistical analyses were performed in Python 3.10.16 using scikit-learn 1.6.1,^28^ SciPy 1.15.2,^29^ XGBoost 3.0.0,^30^ and statsmodels 0.14.4.^31^ A fixed random seed (42) was used to ensure reproducibility.

### Machine Learning Training Pipeline

All machine-learning models were developed using a fully nested cross-validation framework to reduce optimistic bias in model selection and performance estimation.^32^ Participant-level separation was enforced across folds. Because multiple nights of actigraphy were available per participant, the participant identifier was used as the grouping variable during cross-validation. Both outer and inner splits were generated using stratified group cross-validation, ensuring that all nights from a given participant were assigned exclusively to a single fold. Thus, no participant contributed data simultaneously to training and validation or test sets.

The outer loop consisted of 10 stratified group folds. In each iteration, approximately 90% of participants were used for model training, and the remaining 10% were held out as an independent test set. The outer test fold was not accessed during hyperparameter tuning, preprocessing, threshold selection, or early stopping.

Within each outer training fold, hyperparameters were optimized using Optuna^33^, a Bayesian optimization framework that iteratively samples candidate hyperparameter configurations to maximize predefined objective functions. A fixed number of independent trials (n = 250) were evaluated per outer fold, chosen to allow adequate exploration of the hyperparameter space while maintaining computational feasibility. Each trial corresponded to a unique hyperparameter configuration and was assessed using 5-fold stratified group cross-validation restricted to the outer training data. Thus, performance for each configuration reflected the average across inner validation folds. Hyperparameter selection was completed entirely within the outer training folds, and the outer test fold remained unseen until final evaluation.

Thresholds—including the default threshold (*τ* =0.5), the Youden’s J statistic^34^(*τ*\* = argmax_*τ*_(*se*(*τ*) + *sp*(*τ*) - 1)) which gives the best balance between sensitivity and specificity, and constrained thresholds (*τ*_*se*_ or *τ*_*sp*_) selected to maximize sensitivity (Stage 1: questionnaire) or specificity (Stage 2: actigraphy), respectively ^35,36^, were derived exclusively from aggregated predictions generated on inner validation folds. Once selected, thresholds were fixed and applied unchanged to the outer test fold.

After hyperparameter selection, the final model was retrained on the entire outer training set. When early stopping was applied, a validation subset was created solely within the outer training data using stratified group splitting and was not used for performance estimation. Final performance metrics were computed only on the held-out outer test fold.

Because all preprocessing steps (including imputation and scaling) were fitted exclusively on training data within each fold and then applied to validation and test data without refitting, performance estimates reflect fully out-of-sample evaluation at the participant level.

For the questionnaire-based classifier (k = 4 features), we compared Random Forest,^37^ LightGBM,^38^ XGBoost,^30^ and Elastic Net.^39^ Although the questionnaire included only four items, the implementation of ML models enables modeling of potential interaction effects and granular stratifications. The goal was to capture non-linear and combinatorial patterns that might improve discrimination beyond simple additive scoring.

For actigraphy-derived features^18^ (k = 113), we used XGBoost^30^ given its strong performance on high-dimensional tabular data and reduced sensitivity to hyperparameter specification.^40^ Missing values were imputed using feature-wise medians, fit only on training folds.^28^ Feature importance was assessed as regression coefficients for Elastic Net and by the gain metric for tree-based models, where gain quantifies the improvement in model performance attributable to a given split and emphasizes the discriminative value of each feature^30^; Feature importances were aggregated across cross-validation folds, mean and standard deviation were reported.

Performance metrics (sensitivity, specificity, ROC AUC, precision–recall AUC) were aggregated across folds, with 95% confidence intervals estimated using the Student’s t-distribution at *α* =0.05. Performance was reported at *τ, τ*\* and *τ*_*se/sp*_. Adjusted positive predictive value (PPV) was computed at a prevalence of 1.5% in the general population.

Cohort-specific model performance was evaluated using subject-level predictions derived exclusively from outer test folds. For actigraphy, nightly probabilities were averaged within each participant to obtain a single subject-level prediction per fold. Cohort-specific AUCs were computed from these out-of-sample predictions without refitting.

### Two-Stage Screening Approach

Questionnaire and actigraphy data were available for 75 participants with iRBD and 54 controls. To evaluate the proposed two-stage screening framework, we used predictions derived exclusively from outer test folds of the nested cross-validation procedure described above.

For each outer fold, Stage 1 (questionnaire-based) predictions were generated using the model trained on the corresponding outer training fold. Classification thresholds for Stage 1—including the sensitivity-optimized threshold (*τ*_*se*_)—were derived exclusively from inner validation predictions within that outer training fold and were fixed prior to evaluation on the outer test fold.

Stage 2 (actigraphy-based) predictions were similarly generated using actigraphy models trained within each outer training fold. Specificity-optimized thresholds (*τ*_*sp*_) were derived exclusively from inner validation predictions and applied unchanged to the corresponding outer test fold.

The two-stage classification was then constructed using only out-of-sample predictions from the outer test folds. A participant was classified as positive for iRBD only if both the Stage 1 and Stage 2 predictions were positive under their respective predefined thresholds. Participants negative at Stage 1 were not advanced to Stage 2 in the sequential framework.

Importantly, no thresholds were derived using outer test predictions, and no previously published thresholds were reused in this analysis. Final two-stage performance metrics therefore reflect fully out-of-sample, participant-level evaluation.

Sensitivity, specificity, precision, and accuracy were computed by aggregating outer-fold predictions across all participants.

## Results

### Population

Data from 396 participants (99 iRBD, 297 controls) across five cohorts were used in this study. Demographic and clinical characteristics stratified by cohort and diagnosis are detailed in **Table 1**. The pooled sample had a slight male predominance (55.1%).

Compared to other cohorts, participants from VascBrain were overall younger, while the SHAS cohort was the oldest. The Sinai Clinic participants had the highest mean BMI.

Compared with controls, patients with iRBD were generally older, more frequently male, and had lower BMIs. Racial distributions also differed, with a higher proportion of patients with iRBD identifying as White.

### Individual Questions

Group demographic differences (age, BMI, sex) are reported in the Population section in Supplementary Table S1.

The performance of the questionnaire alone was assessed in 95 iRBD and 194 controls. Dream enactment had an AUC of 0.85, sensitivity 77.9%, and specificity 92.3%. Hyposmia had an AUC of 0.69, sensitivity 56.8%, and specificity 80.9%. Constipation had an AUC of 0.62, sensitivity 55.8%, and specificity 67.0%. Orthostatic hypotension had an AUC of 0.52, sensitivity 31.6%, and specificity 75.3% (**Supplementary Table S2**).

In the LASSO regression model, the largest positive coefficients were observed for “dream enactment yes” (β = 0.84) and “smell yes” (β = 0.77). Among interaction terms, the highest coefficients were for interaction between “smell do not know” and “orthostasis yes” (β = 0.65), followed by the interaction between “orthostasis do not know” and “smell yes” (β = 0.58), and the interaction between “orthostasis do not know” and “constipation yes” (β = 0.52). A negative coefficient was identified for the interaction between “dream enactment yes” and “orthostasis do not know” (β = –0.41) (**Supplementary Figure S1**).

### Four-Item Questionnaire

Combining all four questions, XGBoost outperformed all models (**Supplementary Figure S2**). Using Youden’s J statistic, the model achieved an AUC of 0.86 (95% CI: 0.810– 0.910). At the default threshold, the model achieved a sensitivity of 77.9%, and a specificity of 90.7%. At the *τ*_*Se*_ of 0.21, the model achieved a sensitivity of 85.3% and a specificity of 60.8%. At the *r*\* of 0.60, the model achieved a sensitivity of 76.8% and a specificity of 92.3% (**Figure 3**). At an adjusted prevalence of 1.5%, across the folds, the adjusted PPV for the XGBoost model was 10.2 ± 4.0%.

**Figure 3:**
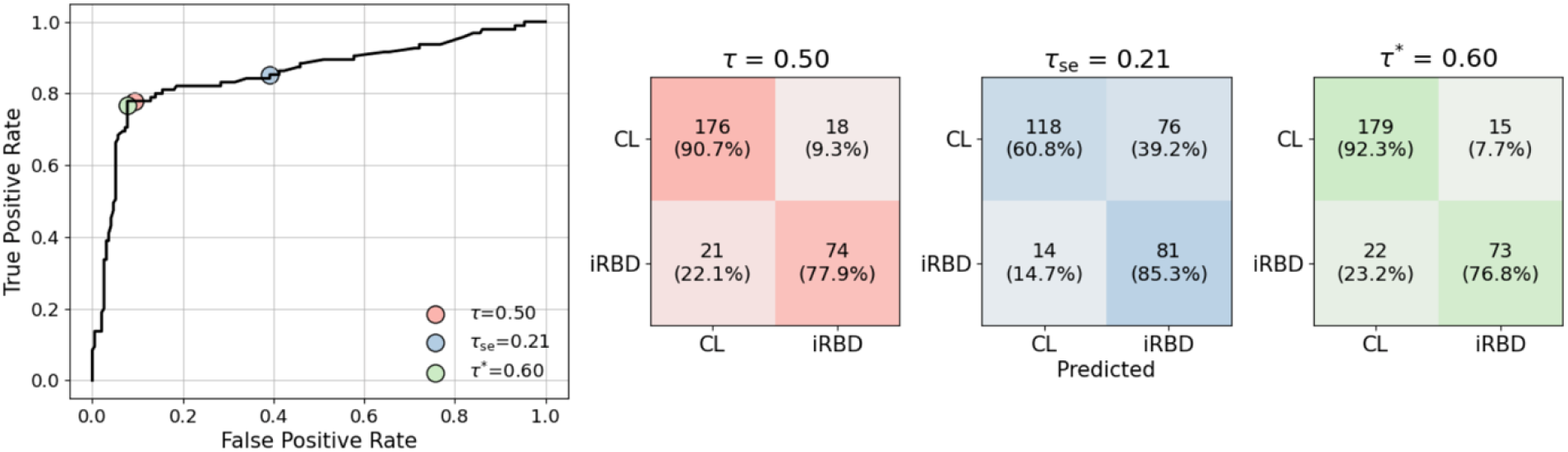
Performance of the XGBoost classifier for questionnaire-based screening. Receiver operating characteristic (ROC) curve for the predicted probabilities with respective matrices corresponding to various thresholds: default (*τ* = 0.50), maximizing sensitivity (*τ*_*Se*_ = 0.21), and Youden’s J (*τ*\* = 0.60). Controls (CL).

Across cohorts, the mean AUC was 0.91 ± 0.10 for the Stanford cohort, 0.86 ± 0.16 for the Sinai cohort, and 0.80 ± 0.25 for the SHAS cohort. On average, validation folds included 50.9% Sinai, 29.1% Stanford, and 20.1% SHAS participants, corresponding to approximately 15, 8, and 6 subjects per fold, respectively.

Feature importance based on average gain across 10 folds identified question dream enactment as the strongest predictor (20.5 ± 15.0), followed by hyposmia (2.3 ± 0.7), constipation (0.8 ± 0.3), and orthostasis (0.6 ± 0.2). XGBoost feature scores across weight, gain, and cover are presented in **Supplemental Figure S3**.

### Actigraphy Screening

Cases with iRBD (N = 78) and controls (N = 158) contributed 1,652 and 2,167 nights of actigraphy data, respectively. Groups were comparable in age and BMI, but the proportion of males was higher among cases with iRBD (70.9% vs. 51.6%, p = 0.0046). Cases with iRBD showed longer total sleep time (7.1 ± 0.9 vs. 6.51 ± 1.04 hours, *p* = 0.0002) and less non-wear hours (0.24 ± 0.32 vs. 0.52 ± 0.46, *p* = 4 × 10-^6^); however, other actigraphy metrics did not differ (**Supplementary Table S3)**.

Based on within-subject probabilities averaged across nights, the XGBoost model achieved an AUC of 0.88 (95% CI: 0.84–0.92) and an area under the precision–recall curve of 0.85 (95% CI: 0.78–0.91). At *r* = 0.5, the model achieved sensitivity 80.8% and specificity 84.8%; at *r*_*Sp*_ = 0.62 sensitivity 61.5% and specificity 96.2%; and at *r*\* = 0.49, sensitivity 82.1% and specificity 83.5%. (**Figure 4**). Performances of the different optimizations are presented in **Supplementary Figure S4**. At an adjusted prevalence of 1.5%, across the folds, the adjusted PPV was 6.2 ± 1.7%. Feature importance is presented in **Supplementary Figure S5**.

**Figure 4:**
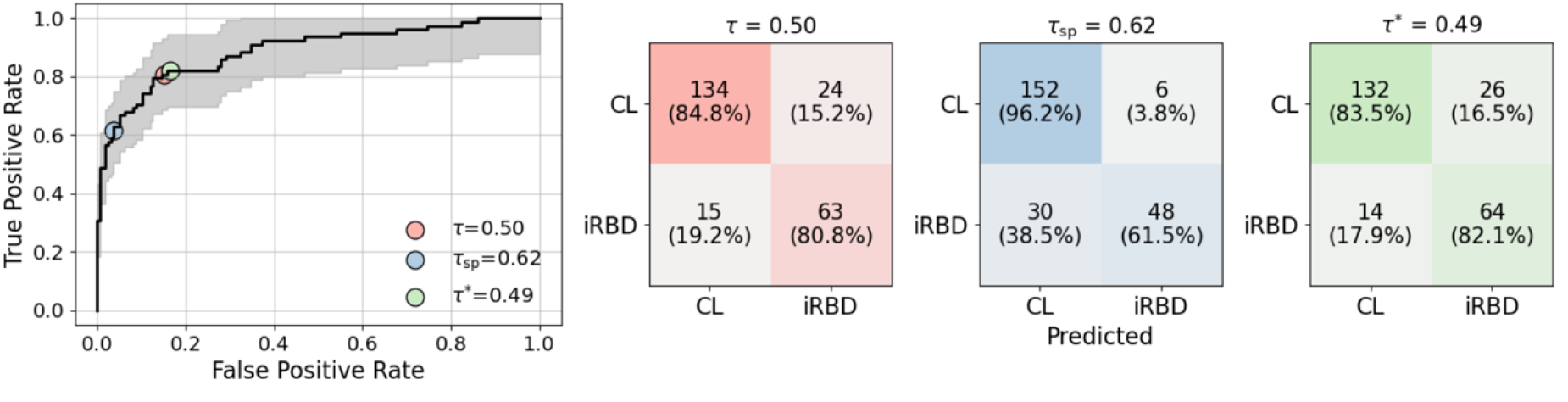
Diagnostic performance of the XGBoost classifier for actigraphy-based screening of 78 cases with iRBD and 158 controls. Receiver operating characteristic curve for the predicted probabilities with thresholds selected by Standard (τ = 0.50), maximum specificity (τ _Sp_= 0.62), and Youden’s J (τ* = 0.49) and their corresponding confusion matrices. Values are presented as counts with row percentages. Controls (CL).

Across cohorts, the mean AUC was 0.98 ± 0.03 for the Stanford cohort, 0.69 ± 0.18 for SHAS, and 1.00 for VascBrain. On average, validation folds included 35.7% for Stanford, 20.8% for SHAS, 10.5% for VascBrain, and 33.0% for ADRC (controls), corresponding to approximately 9, 5, 3, and 8 subjects per fold, respectively.

### Two-Stage Screening

One hundred and twenty-nine participants (iRBD, n = 75; controls, n = 54) completed both questionnaire and actigraphy (see **Supplementary Table S4** for characteristics). Compared to Stanford, the SHAS cohort showed more women and was racially more diverse.

Compared with the actigraphy-only group (n = 107), the two-stage group (n = 129) was younger (65.9 ± 9.1 vs. 67.6 ± 10.2 years; *p* = 0.0501) and predominantly male (69% vs 50.6%, *p*=0.0002) with no significant differences in BMI. Compared with the questionnaire-only group (n = 160), the two-stage group was again older (65.9 ± 9.1 vs 61.15 ± 10.88 years; *p* = 0.0002), had a lower BMI (25.9 ± 4.5 vs. 29.4 ± 6.5; *p* < 4 × 10^-6^), and a higher proportion of White participants (82.7% vs. 51.3%; *p* = 3 × 10^-8^) (**Supplementary Table S5**).

In this cohort, the dream enactment question alone had a sensitivity of 78.4% and specificity of 90.9%, while the four-item questionnaire model had a sensitivity of 87.8% and specificity of 83.6%. The actigraphy model alone had a sensitivity of 79.7% and specificity of 87.3%.

Applied consecutively, the two-stage screening starting with the dream enactment question followed by actigraphy achieved a final sensitivity of 67.6% and specificity of 100%, while using instead the four-item questionnaire (**Figure 5**), final sensitivity and specificity were 68.9% and 100% with default thresholds, and 73.3% and 100%, with optimized thresholds, respectively (**Figure 5**).

**Figure 5:**
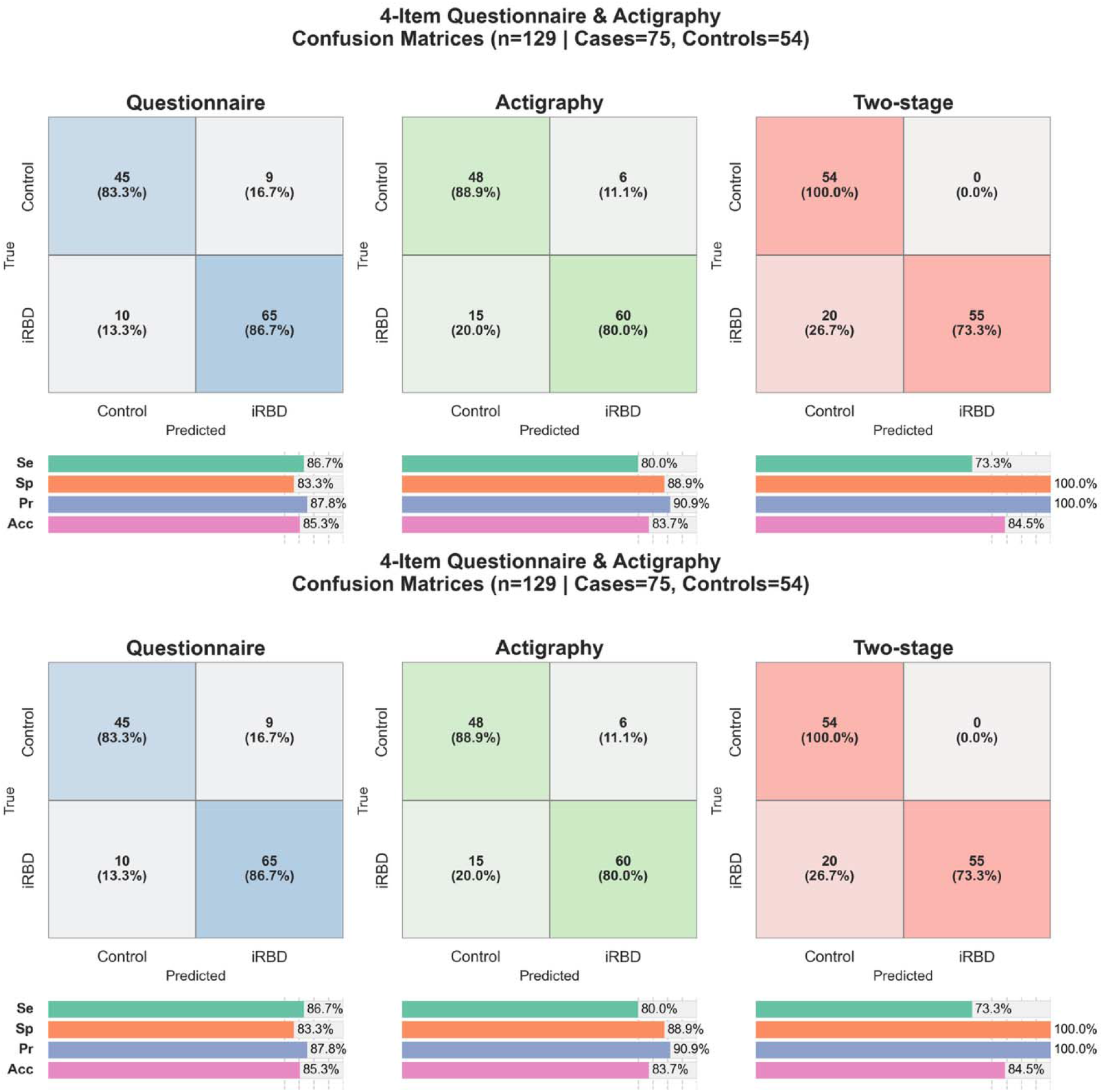
Two-stage screening performance using questionnaire and actigraphy. Confusion matrices and performance metrics are shown for 129 participants with both questionnaire and actigraphy data, applying a sensitivity-optimizing threshold for questionnaire screening and a specificity-optimizing threshold for actigraphy confirmation. (A) Two-stage screening using a single dream enactment question followed by actigraphy. (B) Two-stage screening using a four-item questionnaire followed by actigraphy. Sensitivity (Se); Specificity (Sp); Precision (Pr); Accuracy (Acc).

## Discussion

In this multicenter retrospective case–control study, we evaluated a two-stage screening strategy for identifying iRBD combining a self-administered four-item questionnaire followed by nocturnal wrist actigraphy. Using fully nested cross-validation with participant-level separation, the sequential approach demonstrated strong discrimination in this cohort (sensitivity 73.3%, specificity 100%). Although we recognize that performance estimates from case–control designs may not directly translate to population-based settings, these findings support the feasibility of integrating brief symptom screening with passive wearable monitoring to identify individuals at elevated risk of iRBD.

The first-stage questionnaire screening demonstrated meaningful discrimination despite its brevity. The dream-enactment question was selected based on its previously reported high specificity^41^ relative to other screening instruments^41^, and its performance in the present cohort stresses the clinical relevance of focused symptom inquiry. Machine-learning approaches enabled modeling of potential non-linear and interaction effects among questionnaire items. Compared with the dream-enactment question alone, the four-prodrome model showed a marked increase in sensitivity, from 77.9% to 85.3%. Dream enactment was the most informative single item with 78.4% sensitivity, 90.9% specificity, and an AUC of 0.85. Hyposmia screening showed the second-best performance, with 56.8% sensitivity and 80.9% specificity. Although olfactory loss can be due to other causes, such as anatomical abnormalities and prior COVID-19^41^ infection, it remains strongly associated with Lewy body pathology.^42–44^ Objective testing (e.g., smell kits) is more accurate but adds time, cost, and logistical complexity. In contrast, subjective hyposmia is easy to screen for and reported by about 50% of patients with iRBD^18,45^ and up to 84% with PD^46^. To our knowledge, this is the first study to systematically evaluate the utility of self-reported hyposmia in a large multicenter iRBD cohort.

Actigraphy provided an objective second-stage screen by quantifying nocturnal motor activity patterns characteristic of REM sleep motor dysregulation^41^. The actigraphy classifier achieved an AUC of 0.88 with a sensitivity of 79.7% and specificity of 87.3%, consistent with our recent multicenter validation study (19). The slightly lower performance compared with the previous classifier (AUC = 0.916)^20^ could be due to increased heterogeneity, and greater variability in sleep quality and comorbidities.

The two-stage framework implements a pragmatic screening paradigm: a sensitivity-optimized first stage (questionnaire; *r*_*Se*_) to minimize missed cases, followed by a specificity-optimized second stage (actigraphy; *τ*_*Sp*_) to reduce false positives arising from common clinical mimics of RBD, such as dream-enactment-like behaviors and other abnormal movements due to obstructive sleep apnea, periodic limb movements and non-REM parasomnias. The two-stage strategy yielded a final sensitivity of 73.3% and specificity of 100%. This sequential framework may reduce unnecessary referrals for in-lab sleep testing while maintaining high diagnostic precision across heterogeneous populations.

In contrast, current iRBD screening strategies, which often combine multiple questionnaires followed by expert clinical assessments, have reported lower precision– around 66% in best-case scenarios^14–17^–limiting real-world applicability, especially in population-based screening given the relatively low prevalence of iRBD (∼1.5%). Our approach offers a fully remote, low-cost solution, which could potentially be adapted to consumer wearable ecosystems, pending appropriate validation and ethical oversight.^47^

Strengths of this study include the multicenter design and relatively large sample size. At Mount Sinai, questionnaires were primarily collected in a sleep medicine clinic, contributing to a medically complex cohort with a high rate of symptom endorsement. Questionnaire data did not require an informant and were all collected consecutively, reducing the risk of exclusion bias. All diagnoses of RBD and RBD mimics were confirmed by vPSG. The wearable-based model used explainable features from prior literature, enhancing transparency and interpretability.^21^

This study has several limitations. First, recruitment from established research cohorts and complex sleep cases referred to two academic centers may have enhanced the robustness of the wearable-based algorithm, while potentially limiting sensitivity for detecting milder or subclinical RBD cases outside tertiary referral settings. Similarly, the sensitivity (77.9%) of the self-administered Innsbruck RBD summary question may have been inflated by the inclusion of research participants, generally more aware of and knowledgeable about their diagnosis. A population-based study would be needed to compare the performance of single-question items or extended RBD questionnaires.^48,49^ More broadly, the retrospective case–control design may overestimate diagnostic performance relative to real-world screening settings characterized by lower disease prevalence, greater phenotypic heterogeneity, and less pronounced separation between cases and controls. Second, not all retrospective cohorts followed a harmonized protocol requiring both questionnaire-based screening and wrist actigraphy acquisition. As a result, only a subset of participants had complete data for both stages of the proposed two-stage screening, reducing the effective sample size for final iRBD detection. This partial overlap between datasets may also limit the stability and generalizability of the sequential two-stage performance estimates. Third, reliance on self-reported symptoms, such as dream enactment and hyposmia, introduces the possibility of awareness bias. Individuals with a prior diagnosis of RBD, prior exposure to sleep evaluations, or greater health literacy may be more attuned to these symptoms and therefore more likely to report them, potentially inflating the apparent performance of questionnaire-based screening in case–control settings relative to population-based screening. However, this limitation is partly mitigated by the objective nature of actigraphy in the second stage of the screening framework. Fourth, the influence of comorbid sleep or neurological disorders, medications, and demographic variability on model performance was not systematically assessed. Inter-cohort variability in actigraphy performance further suggests that differences in recruitment source, wearable adherence, and underlying population characteristics may influence the model’s transferability. Finally, because controls who did not endorse dream enactment were not required to obtain bedpartner input or undergo vPSG, some true cases with iRBD may have been mislabeled as controls, resulting in an underestimation of sensitivity.

In conclusion, a two-stage, fully remote screening strategy leveraging brief symptom inquiry and passive actigraphy can identify iRBD with high precision in a multicenter cohort. This approach offers a scalable pathway to enrich at-risk populations for confirmatory testing and trial recruitment, pending prospective validation in community-based samples.

## Acknowledgements

We thank all study participants for enabling this research and the Department of Neurology at the Icahn School of Medicine for funding.

## Author Contributions

CAM, GRM, AM, KHR, EG, FME, ABK, and EHD contributed to the conception and design of the study. CAM, GRM, AM, KHR, SM, LZ, BG, CK, LN, AP, EM, FME, JW, KP, ABK, and EHD contributed to the acquisition and analysis of data. All authors contributed to drafting the text and preparing the figures.

## Data Availability

Anonymized data are available upon reasonable request from any qualified investigator. Code for the analysis is available at: https://github.com/GiorgioRicciardiello/TwoStageRbd.

