## Supplemental Materials for "A Two-Stage Questionnaire and Actigraphy Screening for Isolated REM Sleep Behavior Disorder in a Multicenter Cohort"

### Supplementary

Table S1 -- **Sample characteristics and completed RBD questionnaires**. Responses among participants who completed the RBD questionnaire (N=289), stratified by diagnostic group (RBD vs. controls) and by study cohort (Mount Sinai Clinic, SHAS, Stanford,). Results are reported as mean ± SD (non-missing count) for continuous variables and as counts (percentages) for categorical variables. Statistically significant differences were observed between RBD cases and controls for age (p = 0.0001), BMI (p = 0.0018), gender (p = 0.0203) and race (p = 9 × ${10}^{-5}$). Questionnaire responses also differed significantly between cases and controls for Q1 (RBD symptom; p =1 × ${10}^{-27}$), Q2 (smell loss; p = 5 × ${10}^{-11}$), and Q3 (constipation; p = 0.0002), whereas no significant difference was observed for Q4 (orthostatic symptoms; p = 0.5526).

|  | **Diagnosis** | | | **Cohort** | | |
| --- | --- | --- | --- | --- | --- | --- |
| **variable** | **RBD** | **Control** | **p-value** | **SHAS** | **Sinai** | **Stanford** |
|  | **N=95** | **N=194** |  | **N=58** | **N=147** | **N=84** |
| **Age [years]** | 66.77 ± 8.59 (95) | 61.58 ± 10.77 (194) | 0.0001 | 66.1 ± 9.28 (58) | 60.75 ± 11.05 (147) | 65.79 ± 8.76 (84) |
| **BMI [kg/m²]** | 26.15 ± 4.12 (95) | 28.69 ± 6.48 (193) | 0.0018 | 25.43 ± 4.33 (57) | 29.78 ± 6.56 (147) | 26.12 ± 4.39 (84) |
| **Gender (Male)** | 65 (68.42%) | 105 (54.12%) | 0.0203 | 33 (56.9%) | 75 (51.02%) | 62 (73.81%) |
| **Race** | -- | -- | 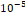 | -- | -- | -- |
| White | 74 (80.43%) | 107 (58.47%) | - | 35 (66.04%) | 69 (50.0%) | 77 (91.67%) |
| Black or African American | 8 (8.7%) | 22 (12.02%) | - | 8 (15.09%) | 21 (15.22%) | 1 (1.19%) |
| Asian | 5 (5.43%) | 9 (4.92%) | - | 2 (3.77%) | 6 (4.35%) | 6 (7.14%) |
| Other | 5 (5.43%) | 43 (23.5%) | - | 7 (13.21%) | 41 (29.71%) | 0 (0.0%) |
| Mixed | 0 (0.0%) | 2 (1.09%) | - | 1 (1.89%) | 1 (0.72%) | 0 (0.0%) |
| **RBD Symptom** | -- | -- | 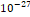 | -- | -- | -- |
| Yes | 74 (77.89%) | 15 (7.73%) | - | 30 (51.72%) | 20 (13.61%) | 39 (46.43%) |
| Do not know | 6 (6.32%) | 41 (21.13%) | - | 7 (12.07%) | 39 (26.53%) | 1 (1.19%) |
| No | 15 (15.79%) | 138 (71.13%) | - | 21 (36.21%) | 88 (59.86%) | 44 (52.38%) |
| **Hyposmia** | -- | -- | 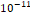 | -- | -- | -- |
| Yes | 44 (46.32%) | 27 (13.92%) | - | 18 (31.03%) | 28 (19.05%) | 25 (29.76%) |
| Do not know | 10 (10.53%) | 10 (5.15%) | - | 7 (12.07%) | 10 (6.8%) | 3 (3.57%) |
| No | 41 (43.16%) | 157 (80.93%) | - | 33 (56.9%) | 109 (74.15%) | 56 (66.67%) |
| **Constipation** | -- | -- | 0.0002 | -- | -- | -- |
| Yes | 47 (49.47%) | 54 (27.84%) | - | 14 (24.14%) | 56 (38.1%) | 31 (36.9%) |
| Do not know | 6 (6.32%) | 10 (5.15%) | - | 8 (13.79%) | 8 (5.44%) | 0 (0.0%) |
| No | 42 (44.21%) | 130 (67.01%) | - | 36 (62.07%) | 83 (56.46%) | 53 (63.1%) |
| **Orthostasis** | -- | -- | 0.5526 | -- | -- | -- |
| Yes | 30 (31.58%) | 48 (24.74%) | - | 13 (22.41%) | 46 (31.29%) | 19 (22.62%) |
| Do not know | 3 (3.16%) | 16 (8.25%) | - | 1 (1.72%) | 17 (11.56%) | 1 (1.19%) |
| No | 62 (65.26%) | 130 (67.01%) | - | 44 (75.86%) | 84 (57.14%) | 64 (76.19%) |

| **Question** | **AUC** | **Sensitivity (%)** | **Specificity (%)** | **Responses** | **Counts** | |
| --- | --- | --- | --- | --- | --- | --- |
|  |  |  |  |  | **RBD**  **(N=**95**)** | **Controls (N=194)** |
| **RBD Symptoms** | 0.85 | 77.9 | 92.3 | Yes | 74 (77.9%) | 15 (7.7%) |
|  |  |  |  | Don’t know | 6 (6.3%) | 41 (21.1%) |
|  |  |  |  | No | 15 (15.8%) | 138 (71.1%) |
| **Hyposmia** | 0.69 | 56.8 | 80.9 | Yes | 44 (46.3%) | 27 (13.9%) |
|  |  |  |  | Don’t know | 10 (10.5%) | 10 (5.2%) |
|  |  |  |  | No | 41 (43.2%) | 157 (80.9%) |
| **Constipation** | 0.62 | 55.8 | 67.0 | Yes | 47 (49.5%) | 54 (27.8%) |
|  |  |  |  | Don’t know | 6 (6.3%) | 10 (5.2%) |
|  |  |  |  | No | 42 (44.2%) | 130 (67.0%) |
| **Orthostatic  hypotension** | 0.52 | 31.6 | 75.3 | Yes | 30 (31.6%) | 48 (24.7%) |
|  |  |  |  | Don’t know | 3 (3.2%) | 16 (8.2%) |
|  |  |  |  | No | 62 (65.3%) | 130 (67.0%) |

**Table S2 –– Predictive performance of individual questionnaire items.** For each question (responses coded as 0 = No, 0.5 = Don’t know, 1 = Yes), AUC was computed, and sensitivity and specificity were derived at the threshold maximizing Youden’s J statistic. Counts and percentages of each response category are shown, stratified by diagnosis; percentages are calculated relative to the total number of cases or controls separately for each question.

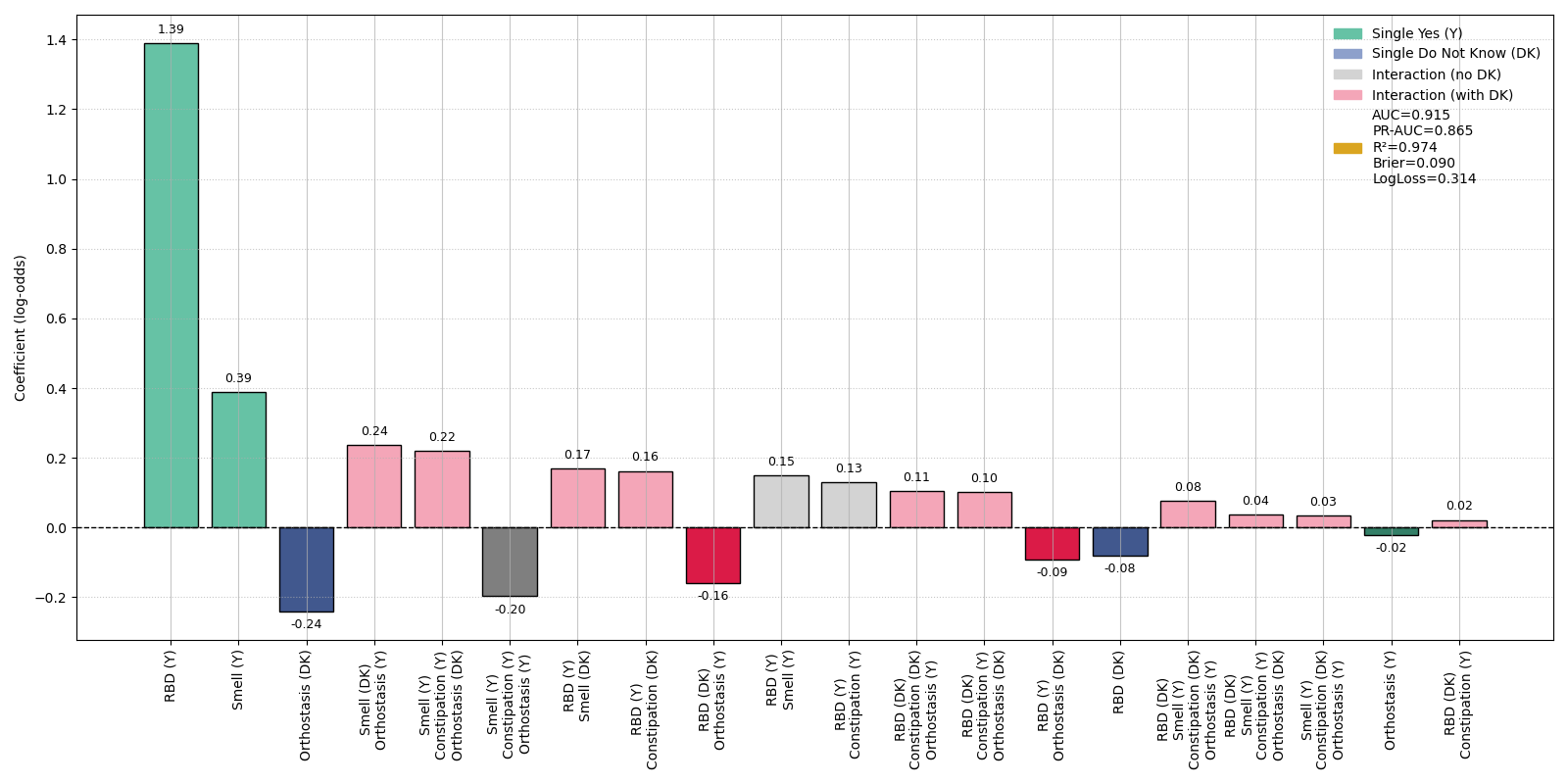

**Figure S1 –– Lasso regression coefficients for four-item questionnaire responses used in RBD classification.** Positive coefficients indicate features contributing to higher predicted probability of RBD, with the strongest effects observed for RBD “yes” (β = 1.39) and smell “yes” (β = 0.39). Interaction terms contributed substantially, particularly “smell do not know” × “orthostasis yes” (β = 0.24), “orthostasis do not know” × “smell yes” (β = 0.22), and “RBD yes” × “smell yes” (β = 0.17). Negative coefficients were observed for orthostasis “do not know” (β = –0.24), “smell no” × “constipation yes” (β = –0.20), and “RBD yes” × “orthostasis do not know” (β = –0.16). Model performance is shown on the right (AUC = 0.915, PR-AUC = 0.865, R² = 0.974, Brier score = 0.090, log-loss = 0.314) evaluated in the full dataset.

| 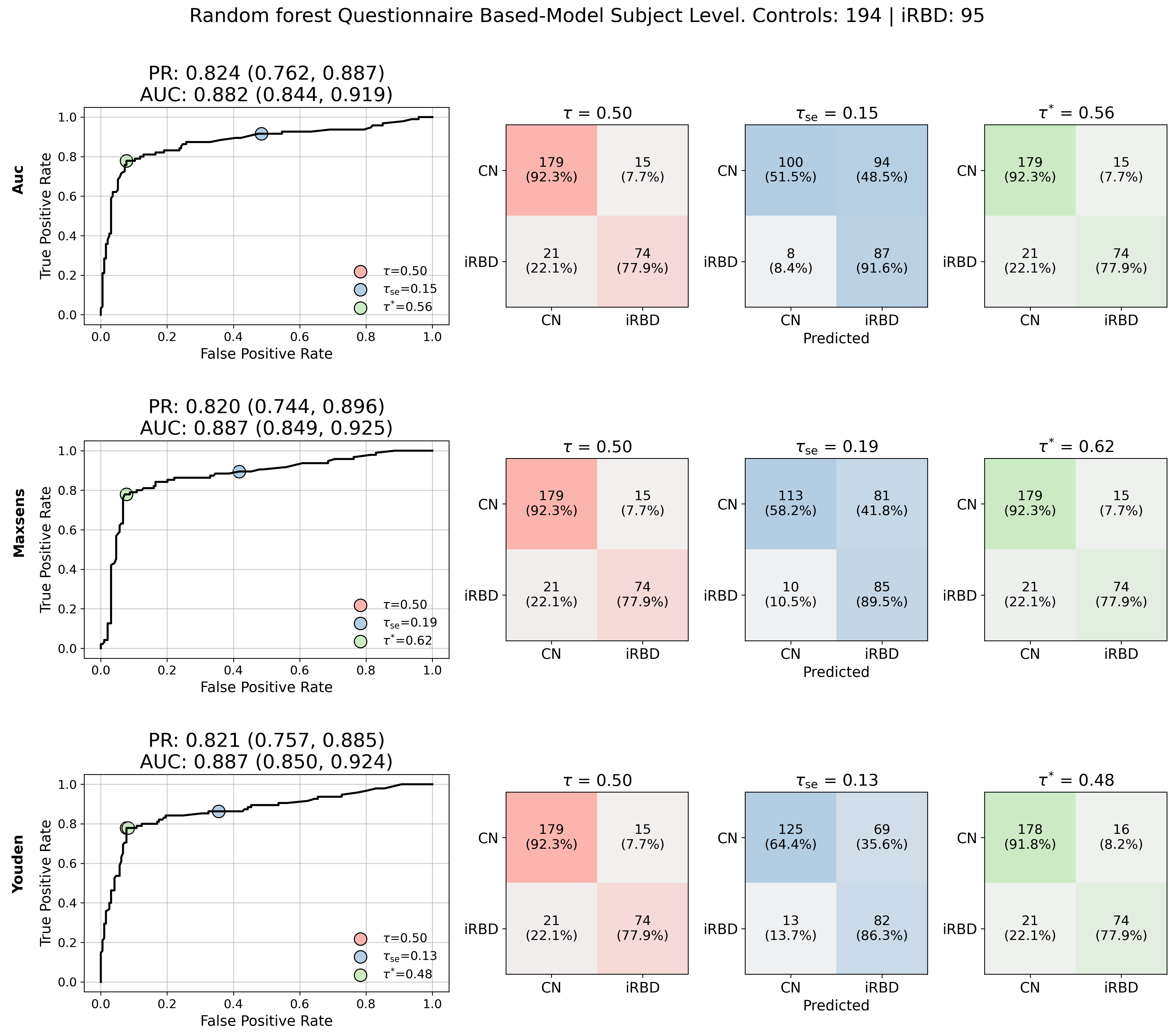  **A – Random Forest** | 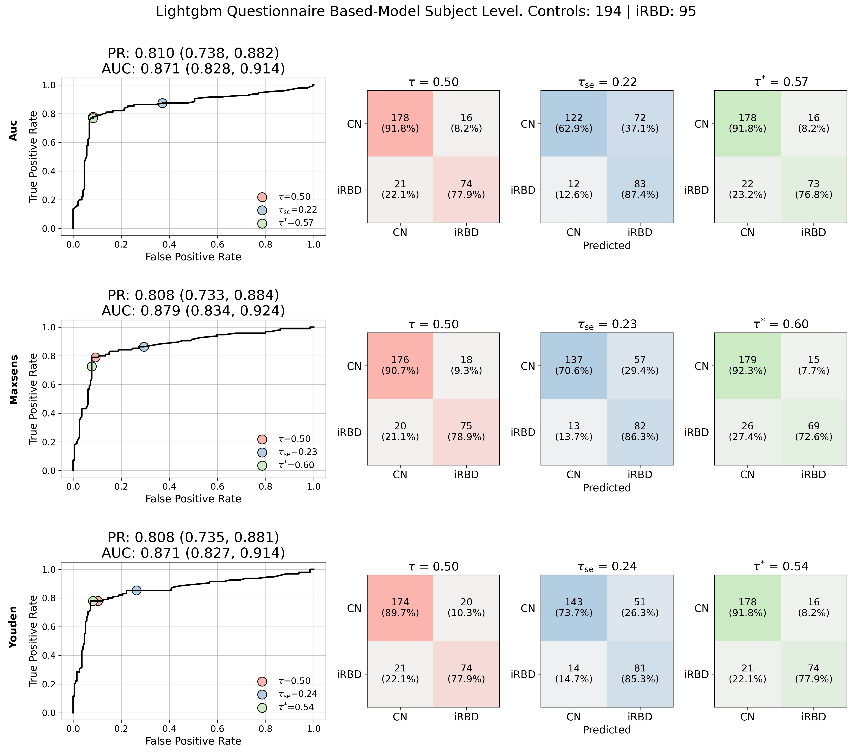  **B – Light GBM** |
| --- | --- |
| 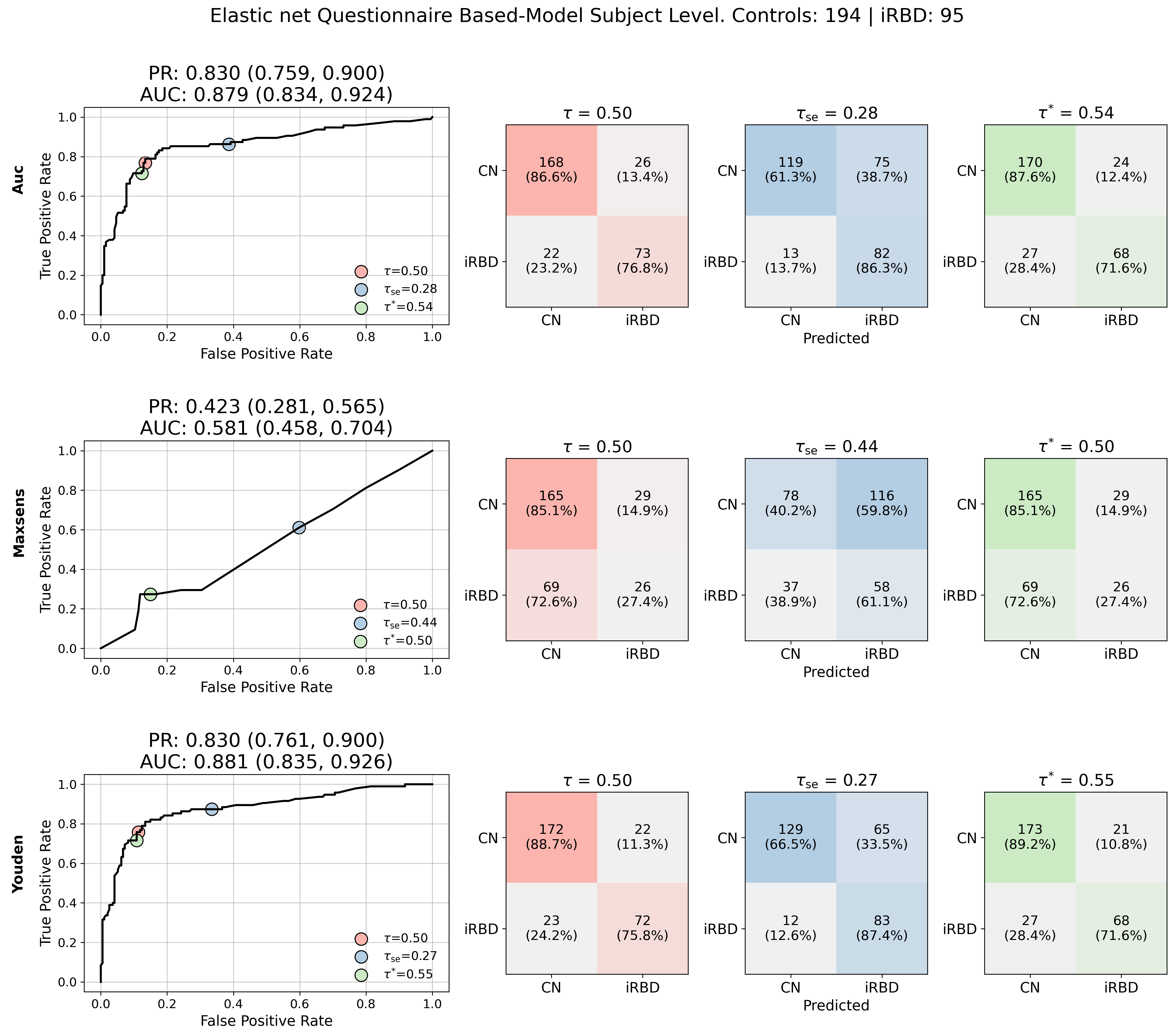  **C – Elastic Net** | 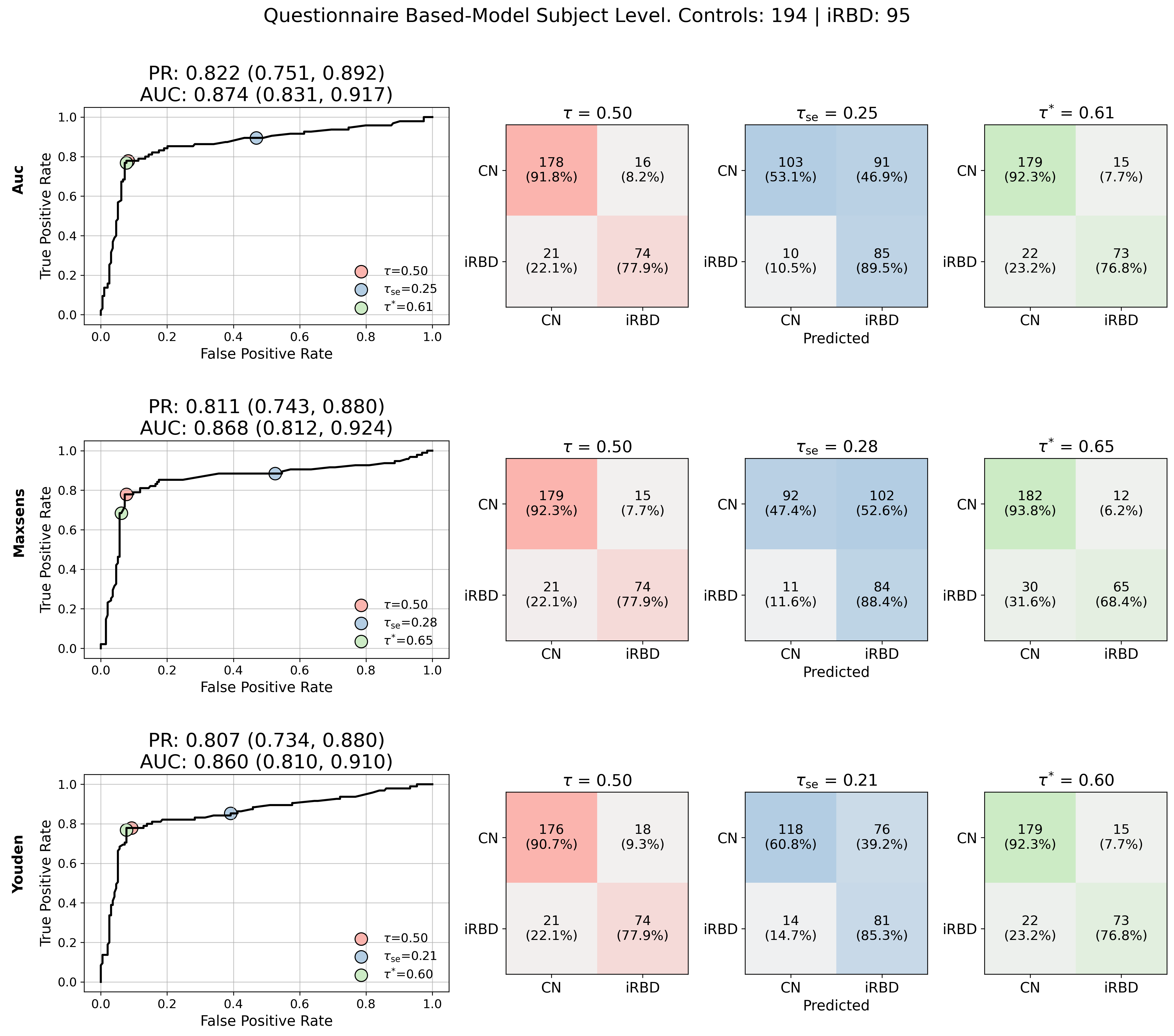  **D - XGBoost** |

Figure S2 –– **Performance of the classifier models under different optimizations** **for questionnaire-based model.** A) Random Forest, B) Light GBM, C) Elastic Net, and D) XGBoost. Each model performance presents a grid of ROC curves and confusion matrices. Each row represents a different optimization strategy (AUC, maximum sensitivity, and Youden’s J statistic), and each column corresponds to a classification threshold ($\tau=$0.5, $\tau^{*}$ and $\tau_{se/sp}$). Confusion matrices display counts and percentages for the corresponding thresholds indicating sensitivity and specificity. Reported AUC and precision-recall curve include 95% confidence intervals. Model criteria were selected as the best trade-off between sensitivity and specificity and the AUC score.

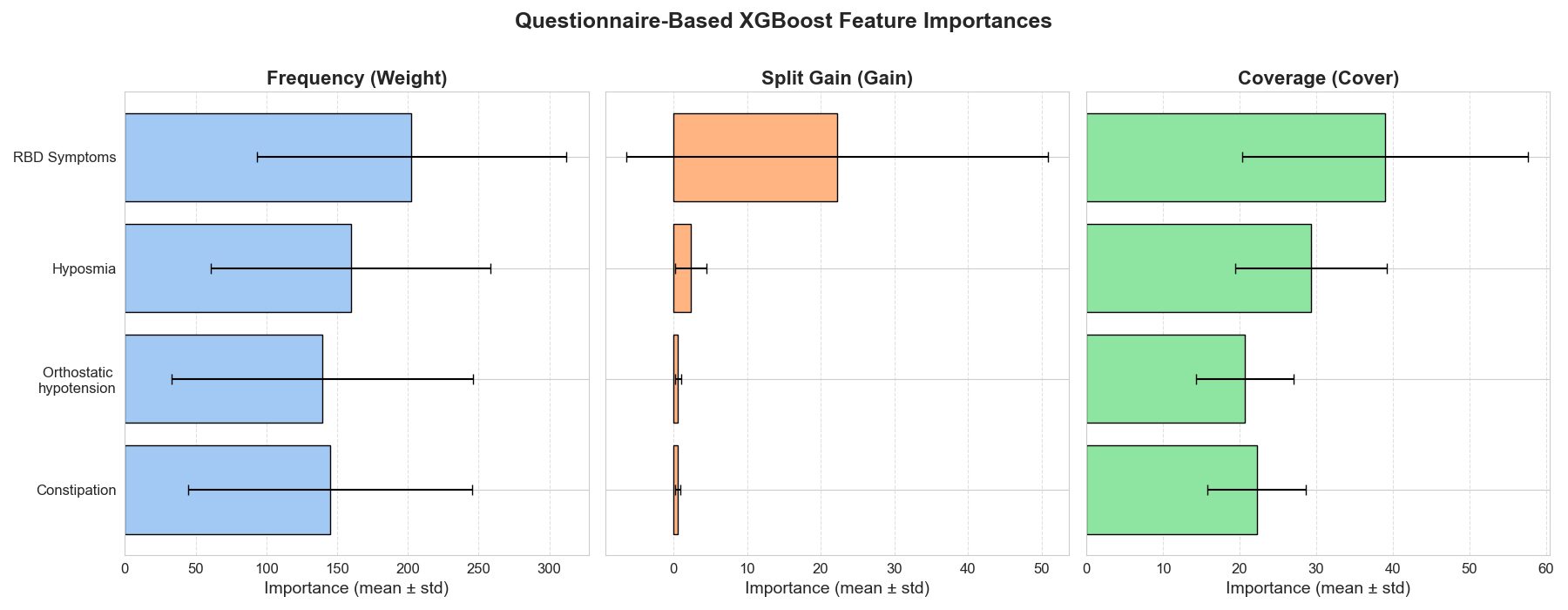

Figure S3 -- **Questionnaire based feature importance for the XGBoost model.** Feature importance across 10 outer folds for the questionnaire-based model. Feature importance was quantified using three complementary metrics: frequency (weight), average improvement in predictive performance per split (gain), and average sample coverage per split (cover). Across all metrics, RBD Symptoms consistently emerged as the most influential feature, with additional contributions from Hyposmia, Constipation, and Orthostatic hypotension. Bars represent the mean importance across folds, and error bars denote the standard deviation from the average across the 10 folds.

Table S3 -- **Actigraphy Screening sample distribution**. Sample characteristics and objective sleep measures from actigraphy (total sleep time (TST), wake after sleep onset (WASO), and sleep efficiency (SE). Distributions are stratified by diagnosis (RBD vs controls) and by cohort (SHAS, Stanford, VascBrain).

| **Diagnosis** | | | | |
| --- | --- | --- | --- | --- |
| **variable** | **Control N=158** | **RBD N=78** | **p-value** | |
| **Age [years]** | 66.5 ± 10.3 (157) | 67.0 ± 8.3 (78) | 0.8026 | |
| **BMI [kg/m²]** | 25.9 ± 4.8 (67) | 26.3 ± 4.2 (78) | 0.6431 | |
| **Gender (Male)** | 81 (51.59%) | 56 (70.89%) | 0.0046 | |
| **Race** | -- | -- | 0.4143 | |
| White | 133 (86.36%) | 59 (80.82%) | - | |
| Black or African American | 1 (0.65%) | 7 (9.59%) | - | |
| Asian | 11 (7.14%) | 4 (5.48%) | - | |
| Other | 5 (3.25%) | 3 (4.11%) | - | |
| Mixed | 4 (2.6%) | 0 (0.0%) |  |  |
| **TST** | 6.9 ± 1.1(158) | 7.1 ± 0.9 (78) | 0.3057 | |
| **WASO** | 1.7 ± 0.6(158) | 1.8 ± 0.7 (78) | 0.3271 | |
| **SE** | 0.8 ± 0.1(158) | 0.8 ± 0.1 (78) | 0.9099 | |
| **Cohort** | | | | |
| **variable** | **SHAS N=49** | **Stanford N=84** | **ADRC N=78** | **VascBrain N=25** |
| **Age [years]** | 65.8 ± 1.0 (48) | 65.8 ± 8.8 (84) | 71.7 ± 6.9 (78) | 55.7 ± 9.1 (25) |
| **BMI [kg/m²]** | 25.9 ± 4.9 (48) | 26.1 ± 4.4 (84) | - | 26.8 ± 4.0 (14) |
| **Gender (Male)** | 31 (63.27%) | 62 (73.81%) | 34 (43.59%) | 10 (40.0%) |
| **Race** | -- | -- | -- | -- |
| White | 28 (65.12%) | 77 (91.67%) | 68 (87.18%) | 19 (86.36%) |
| Black or African American | 6 (13.95%) | 1 (1.19%) | 1 (1.28%) | 0 (0.0%) |
| Asian | 2 (4.65%) | 6 (7.14%) | 7 (8.97%) | 0 (0.0%) |
| Other | 6 (13.95%) | 0 (0.0%) | 1 (1.28%) | 1 (4.55%) |
| Mixed | 1 (2.33%) | 0 (0.0%) | 1 (1.28%) | 2 (9.09%) |
| **TST** | 6.7 ± 1.3 (49) | 6.86 ± 0.8 (84) | 7.3 ± 1 (78) | 6.52 ± 1.1 (25) |
| **WASO** | 2.0 ± 0.7 (49) | 1.71 ± 0.6 (84) | 1.55 ± 0.6 (78) | 2.11 ± 0.5 (25) |
| **SE** | 0.77 ± 0.1 (49) | 0.8 ± 0.1 (84) | 0.82 ± 0.07 (78) | 0.76 ± 0.1 (25) |

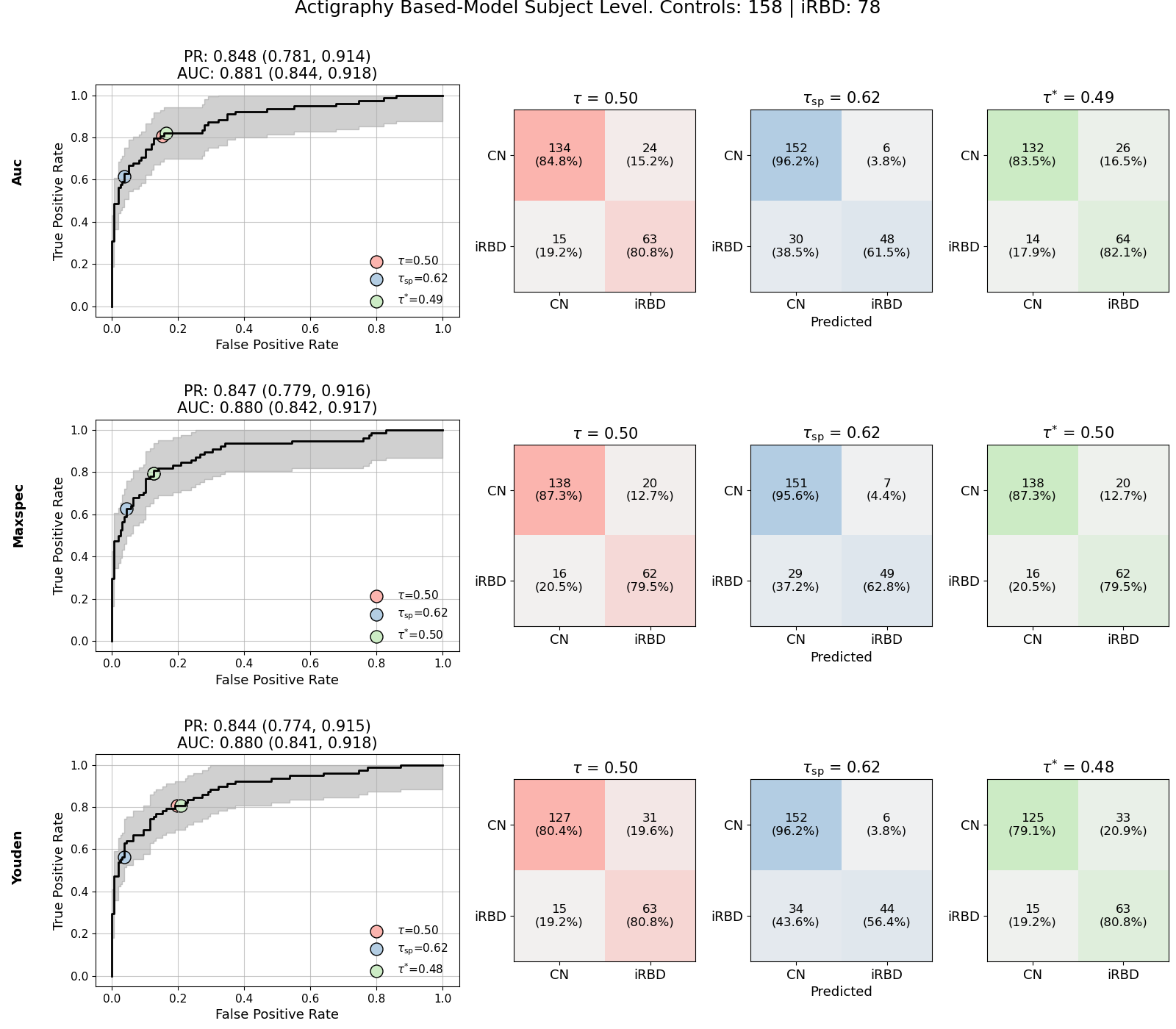

Figure S4 -- **Performance of the XGBoost model under different optimizations** **for actigraphy-based model.** Grid of ROC curves and confusion matrices for the actigraphy-based XGBoost model, trained with nested cross-validation and Optuna-based hyperparameter tuning. Each row represents a different optimization strategy (AUC, maximum sensitivity, or Youden’s J statistic), and each column corresponds to a classification threshold ($\tau^{*}$, fixed 0.5, or $\tau_{sp}$). ROC plots show mean curves with standard deviation across 10 outer folds, while confusion matrices display counts and percentages for the corresponding thresholds. Reported metrics (AUC, sensitivity, specificity) include 95% confidence intervals The Youden-optimized model achieved the best balance between sensitivity and specificity, while the sensitivity-maximized model prioritized case detection at the cost of increased false positives.

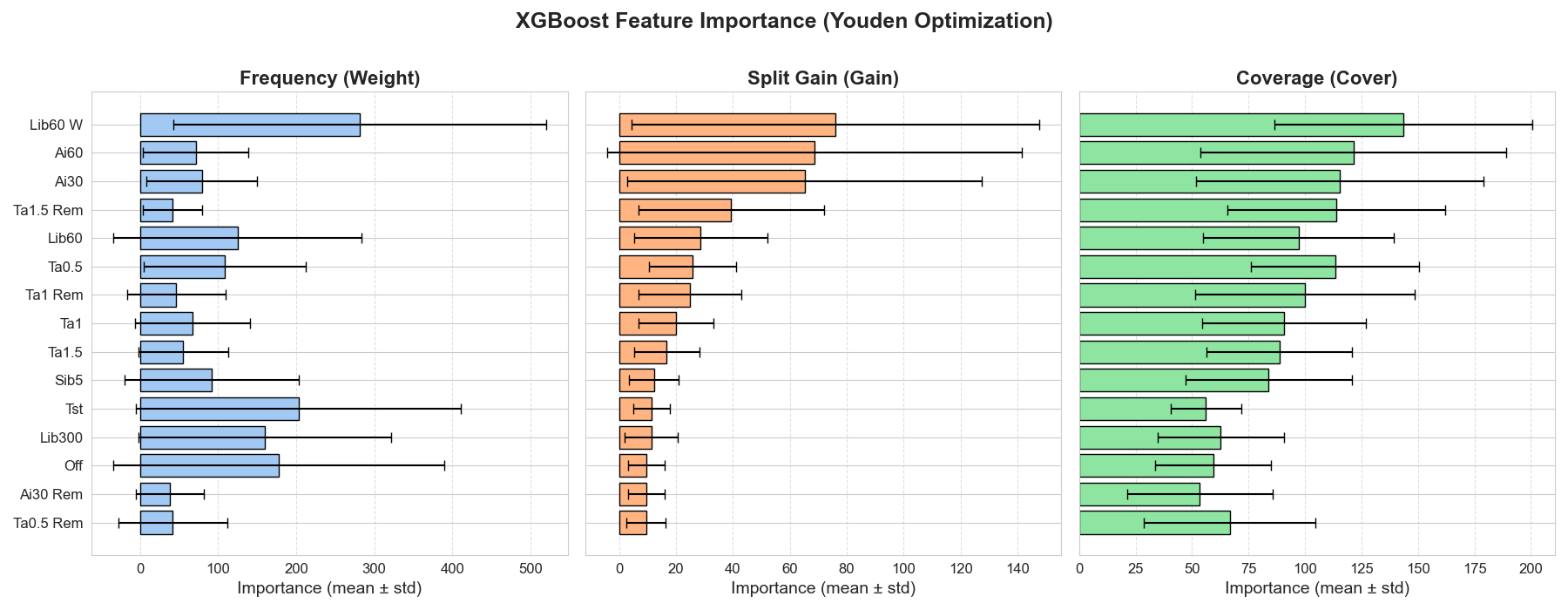

Figure S5 **-- XGBoost feature importance under Youden optimization across outer folds for the actigraphy-based model.** Feature importance was evaluated using three complementary metrics: frequency (weight), average gain in predictive performance per split (gain), and average sample coverage per split (cover). Bars represent the mean importance across folds, and error bars denote the standard deviation computed across the outer folds. The analysis highlights Lib60 W and Ai60 as the most influential features by gain, while other parameters such as Ai30, Ta0.5 Rem, and Lib60 contributed substantially.

Table S4 -- **Two-stage screening sample distribution**. Sample distribution for 75 RBD and 54 controls completed both the questionnaire and actigraphy measures. Distribution of the subset is stratified by diagnosis and by cohort, respectively. Reported values include mean ± SD (with sample size) for continuous variables and counts (percentages) for categorical variables. P-values correspond to between-group statistical tests.

|  | **Diagnosis** | | | **Cohort** | | |
| --- | --- | --- | --- | --- | --- | --- |
| **variable** | **RBD N=75** | **Control N=54** | **p-value** | **SHAS N=45** | **Stanford N=84** | **p-value** |
| **Age [years]** | 67.3 ± 8.1 (75) | 64.1 ± 10.1 (54) | 0.0775 | 66.2 ± 9.8 (45) | 65.8 ± 8.8 (84) | 0.5847 |
| **BMI [kg/m²]** | 26.1 ± 4.1 (75) | 25.7 ± 5.0 (54) | 0.5879 | 25.6 ± 4.6 (45) | 26.1 ± 4.4 (84) | 0.4050 |
| **Gender (Male)** | 52 (69.33%) | 37 (68.52%) | 0.9214 | 27 (60.0%) | 62 (73.81%) | - |
| **Race** | -- | -- | 0.6895 | -- | -- | 0.0002 |
| White | 59 (80.82%) | 46 (85.19%) | - | 28 (65.12%) | 77 (91.67%) | - |
| Black or African American | 7 (9.59%) | 0 (0.0%) | - | 6 (13.95%) | 1 (1.19%) | - |
| Asian | 4 (5.48%) | 4 (7.41%) | - | 2 (4.65%) | 6 (7.14%) | - |
| Other | 3 (4.11%) | 3 (5.56%) | - | 6 (13.95%) | 0 (0.0%) | - |
| Mixed | 0 (0.0%) | 1 (1.85%) | - | 1 (2.33%) | 0 (0.0%) | - |
| TST | 7.04 ± 0.83 (74) | 6.52 ± 1.01 (54) | 0.0025 | 6.76 ± 1.18 (45) | 6.86 ± 0.79 (84) | 0.8744 |
| WASO | 1.8 ± 0.66 (74) | 1.82 ± 0.62 (54) | 0.7874 | 2.0 ± 0.71 (45) | 1.71 ± 0.58 (84) | 0.0811 |
| SE | 0.8 ± 0.07 (74) | 0.79 ± 0.07 (54) | 0.1714 | 0.78 ± 0.08 (45) | 0.8 ± 0.06 (84) | 0.1004 |
| **RBD Symptom** | -- | -- | 6 × ${10}^{-15}$ | -- | -- | 0.1565 |
| Yes | 58 (77.33%) | 5 (9.26%) | - | 24 (53.33%) | 39 (46.43%) | - |
| Do not know | 4 (5.33%) | 3 (5.56%) | - | 6 (13.33%) | 1 (1.19%) | - |
| No | 13 (17.33%) | 46 (85.19%) | - | 15 (33.33%) | 44 (52.38%) | - |
| **Hyposmia** | -- | -- | 2 × ${10}^{-8}$ | -- | -- | 0.5679 |
| Yes | 33 (44.0%) | 4 (7.41%) | - | 12 (26.67%) | 25 (29.76%) | - |
| Do not know | 10 (13.33%) | 0 (0.0%) | - | 7 (15.56%) | 3 (3.57%) | - |
| No | 32 (42.67%) | 50 (92.59%) | - | 26 (57.78%) | 56 (66.67%) | - |
| **Constipation** | -- | -- | 6 × ${10}^{-7}$ | -- | -- | 0.7068 |
| Yes | 36 (48.0%) | 5 (9.26%) | - | 10 (22.22%) | 31 (36.9%) | - |
| Do not know | 6 (8.0%) | 2 (3.7%) | - | 8 (17.78%) | 0 (0.0%) | - |
| No | 33 (44.0%) | 47 (87.04%) | - | 27 (60.0%) | 53 (63.1%) | - |
| **Orthostasis** | -- | -- | 0.0140 | -- | -- | 0.9097 |
| Yes | 23 (30.67%) | 7 (12.96%) | - | 11 (24.44%) | 19 (22.62%) | - |
| Do not know | 1 (1.33%) | 0 (0.0%) | - | 0 (0.0%) | 1 (1.19%) | - |
| No | 51 (68.0%) | 47 (87.04%) | - | 34 (75.56%) | 64 (76.19%) | - |

**Table S5 -- Demographic and clinical characteristics of participants stratified by assessment group**. Comparisons are shown between the two-stage group (questionnaire + actigraphy, N = 129) against the questionnaire-only group (N = 160), and the two-stage groups against actigraphy-only group (N = 107). Reported values include mean ± SD (with sample size) for continuous variables and counts (percentages) for categorical variables. P-values correspond to between-group statistical tests.

| **variable** | **Two Stage (N=129)** | **Questionnaire Only (N=160)** | **p-value** | **Actigraphy Only (N=107)** | **p-value** |
| --- | --- | --- | --- | --- | --- |
| **Age [years]** | 65.94 ± 9.1 (129) | 61.15 ± 10.88 (160) | 0.0002 | 67.56 ± 10.23 (107) | 0.0501 |
| **BMI [kg/m²]** | 25.95 ± 4.47 (129) | 29.4 ± 6.49 (159) | 4 × ${10}^{-6}$ | 27.42 ± 4.6 (17) | 0.1727 |
| **Gender (Male)** | 89 (68.99%) | 81 (50.62%) | 0.0016 | 48 (44.86%) | 0.0002 |
| **Race** | -- | -- | 3 × ${10}^{-8}$ | -- | 0.4443 |
| White | 105 (82.68%) | 76 (51.35%) | - | 87 (87.0%) | - |
| Black or African American | 7 (5.51%) | 23 (15.54%) | - | 1 (1.0%) | - |
| Asian | 8 (6.3%) | 6 (4.05%) | - | 7 (7.0%) | - |
| Other | 6 (4.72%) | 42 (28.38%) | - | 2 (2.0%) | - |
| Mixed | 1 (0.79%) | 1 (0.68%) | - | 3 (3.0%) | - |
| **RBD Symptom** | -- | -- | 8 × ${10}^{-5}$ | - | - |
| Yes | 63 (48.84%) | 26 (16.25%) | - | - | - |
| Do not know | 7 (5.43%) | 40 (25.0%) | - | - | - |
| No | 59 (45.74%) | 94 (58.75%) | - | - | - |
| **Hyposmia** | -- | -- | 0.1035 | -- | - |
| Yes | 37 (28.68%) | 34 (21.25%) | - | - | - |
| Do not know | 10 (7.75%) | 10 (6.25%) | - | - | - |
| No | 82 (63.57%) | 116 (72.5%) | - | - | - |
| **Constipation** | -- | -- | 0.3748 | -- | - |
| Yes | 41 (31.78%) | 60 (37.5%) | - | - | - |
| Do not know | 8 (6.2%) | 8 (5.0%) | - | - | - |
| No | 80 (62.02%) | 92 (57.5%) | - | - | - |
| **Orthostasis** | -- | -- | 0.0089 | -- | - |
| Yes | 30 (23.26%) | 48 (30.0%) | - | - | - |
| Do not know | 1 (0.78%) | 18 (11.25%) | - | - | - |
| No | 98 (75.97%) | 94 (58.75%) | - | - | - |
| **Diagnosis (RBD)** | 54 (41.86%) | 140 (87.5%) | 2 × ${10}^{-16}$ | 103 (96.26%) | 1 × ${10}^{-18}$ |
